# Randomized metformin and cognitive outcomes in the Diabetes Prevention Program Outcomes Study

**DOI:** 10.64898/2026.08.05.26359234

**Authors:** Pandora L. Wander, Lindsay Doherty, Qing Pan, Owen Carmichael, Raymond Turner, Shihchen Kuo, Medha Munshi, Amisha Wallia, James M. Noble, Vallabh O. Shah, Neelesh K. Nadkarni, Sunder Mudaliar, Dana Dabelea, Marinella Temprosa, William C. Knowler, David M. Nathan, José A. Luchsinger, the DPP Research Group

## Abstract

**Importance:** Metformin may influence risk of dementia, with prior conflicting observations of protection or harm.

**Objective:** To determine the association of randomization to metformin vs. placebo or intensive lifestyle intervention (ILS) in the Diabetes Prevention Program (DPP) with cognitive outcomes (cognitive impairment syndromes and trajectories of cognitive test performance) during the DPP Outcomes Study (DPPOS).

**Design, Setting & Participants:** Prospective long-term follow-up of DPP/DPPOS participants at 27 U.S. centers among adults who were at high risk for type 2 diabetes (T2D) at baseline.

**Exposures:** Randomization to metformin, placebo, or ILS (1996–1999) for 3.2 years followed by open-label metformin in the original randomized metformin group until 2021.

**Main Outcomes & Measures:** Cognitive impairment syndromes were adjudicated in 2022– 2024 in 1,483 participants (median age 74 [IQR 68, 80]) using the National Alzheimer’s Coordinating Center Uniform Dataset version 3. Cognitive performance in executive and memory domains was ascertained with repeated cognitive tests between 2009 and 2024. Multinomial logistic regression and mixed-effects models were fit to examine associations of randomization to metformin with cognitive outcomes.

**Results:** Total metformin exposure (mean ± SD) was 15.5 ±7.7 years/person in the metformin group. Persons in the placebo and ILS groups received out-of-study metformin usually after developing diabetes with mean metformin total exposure of 4.5 ±5.1 and 3.8 ±4.8 years/person in the placebo and ILS groups, respectively. Overall, the frequency distributions of the cognitive syndromes did not differ significantly by treatment group; however, randomization to metformin was associated with a 60% (OR 0.40 [95%CI 0.17, 0.97]) and 62% (OR 0.38 [95%CI 0.16, 0.89]) lower odds of dementia compared with placebo and ILS, respectively, after adjustment for demographics, education, income, and APOE-ε4 genotype. Randomization to metformin was also associated with significantly better memory performance over time (β=0.58; 95%CI: 0.09, 1.1; p=0.02; Cohen’s d=0.1).

**Conclusions and Relevance:** Long-term metformin treatment is associated with a reduced risk of dementia and better memory performance among persons with pre-diabetes or T2D. Estimates were imprecise due to a limited number of dementia cases. Longer follow-up with more dementia cases is needed to confirm our findings.

**KEY POINTS:** *Question:* Is chronic metformin treatment related to the risk of dementia and cognitive impairment?

*Findings:* Randomization to metformin in the Diabetes Prevention Program was associated with a lower risk of dementia in the Diabetes Prevention Program Outcomes Study compared with the randomization to placebo or randomization to intensive lifestyle intervention, but the overall distribution of cognitive impairment syndromes did not differ significantly by treatment group. Randomization to metformin was also related to modestly better longitudinal performance in a memory test.

*Meaning:* Chronic metformin treatment may decrease the risk of dementia.

## INTRODUCTION

Metformin is the traditional first-line and most-used medicine for type 2 diabetes (T2D)^1^ and is used off-label for polyendocrine metabolic ovarian syndrome^2^ and prediabetes.^3^ In the three-year Diabetes Prevention Program (DPP), metformin decreased the incidence of T2D in adults at risk by 31% compared with placebo.^3^ Following the masked DPP treatment period (1996– 2001), the program transitioned to a largely observational follow-up study, the DPP Outcomes Study (DPPOS, 2002–present). During DPPOS, persons in the original randomized metformin group transitioned to open-label metformin, which was discontinued on Jan. 31, 2021.^4^ The beneficial effects of metformin on glycemia and T2D risk persisted at 10 years^5^ and 20 years^6^ following conclusion of the randomized trial.

Metformin has pleiotropic effects beyond glucose lowering and has hypothesized anti-aging benefits.^7^ The effects of metformin on cognition are controversial,^8^ with different epidemiologic studies suggesting that metformin increases^9,10^ or decreases^11^ the risk of dementia. Meta-analyses of observational studies suggest that chronic metformin use is associated with a lower risk of neurodegenerative disorders including Alzheimer’s disease (AD) among persons with diabetes.^12^ However, observational studies are limited by the use of administrative datasets, claims data, or lack of control for confounding by indication.^8^ Randomized clinical trials of metformin on cognitive outcomes are limited to small short-term studies.^13,14^

The design of DPP/DPPOS overcomes many of these limitations with more than two decades of exposure to randomized metformin and late-life comprehensive cognitive assessments which allow for determination of cognitive clinical syndromes. We examined the association of randomization to metformin with more than 10 years of cognitive testing including rigorous ascertainment of cognitive impairment syndromes including dementia in 2022–2024. We hypothesized that original randomization to metformin in the DPP followed by open label metformin would be associated with lower risk of cognitive impairment including dementia and with slower cognitive decline.

## METHODS

### Study design, population, and participants

We compared cognitive outcomes in participants randomly assigned to masked metformin during DPP in 1996–1999 to those in participants assigned to placebo or intensive lifestyle intervention (ILS). Cognitive impairment syndromes were determined at a single exam during DPPOS (2022–2024) while serial cognitive testing was conducted five times beginning in 2009– 2010. The eligibility criteria, design, and methods of the DPP^3^ and DPPOS^5^ have been reported elsewhere. Briefly, the DPP was a randomized clinical trial comparing the effects of ILS, masked metformin (850 mg twice daily), and placebo on diabetes incidence among 3,234 adults at high risk for type 2 diabetes. At entry (1996–1999), participants were required to have BMI ≥24 kg/m² (≥22 kg/m² in Asian Americans), fasting plasma glucose levels 95–125 mg/dl (no lower limit in the American Indian clinics), and impaired glucose tolerance (2-hour post-load glucose of 140–199 mg/dl). Participants were excluded if taking medicines known to alter glycemia or if they had illnesses that could reduce their life expectancy or ability to participate in the trial. Masked treatment was discontinued in July 2001, after an average intervention duration of 3.2 years.^5^ Beginning in 2002 and following a 13-month bridge period, 2,779 DPP participants (86%) continued follow-up in DPPOS. All DPPOS participants were offered group-implemented lifestyle intervention, and the original metformin group participants were offered an open-label extension of metformin treatment that continued until 2021. The current analyses are limited to 1,483 DPP/DPPOS participants who completed sufficient cognitive assessment for clinical classification between 2022 and 2024 (**Supplemental Table 1, Supplemental Figure 1**). All participants provided written informed consent.

### Exposure assessment

The main exposure variable was randomization to metformin in the DPP, following an intention-to-treat (ITT) principle. The ITT analysis does not consider out-of-study metformin, which was prescribed by the participants’ clinicians after diabetes developed. We previously demonstrated that inclusion of post-trial, nonrandomized metformin exposure may lead to biases compared to the ITT estimates, most importantly confounding by indication and differential treatment crossover.^15^ Randomization to metformin included a double-blinded phase between 1996 and 2001, followed by an open-label phase in participants originally randomized to metformin until January 31, 2021. Persons originally randomized to metformin had a mean ± standard deviation exposure of 15.5 ±7.7 years of total metformin exposure (study and out-of-study) vs. 4.5 ±5.1 years and 3.8 ±4.8 years of out-of-study metformin exposure in the placebo and ILS groups, respectively. **Supplemental Figure 2** displays total and out-of-study patterns of metformin use over time by treatment assignment.

### Outcome Measures

The primary cognitive outcomes were clinically significant cognitive impairment syndromes used in research and clinical practice including dementia. Cognitive syndromes were assessed during the 2022–2024 DPPOS study visit at which all participants were invited to complete National Alzheimer’s Coordinating Center uniform dataset version 3 (NACC UDSv3) neuropsychological test battery,^16^ along with informant-reported functional and cognitive performance.^17^ Clinic staff administered the NACC UDSv3 neuropsychological battery that included the Montreal Cognitive Assessment (MOCA), Craft Story 21 Recall test, trail making tests (TMT) A and B, Benson visual retention test, Digit Span Forward and Backward, category fluency for animals and vegetables, letter fluency, and the multilingual naming test (MINT).^16^ A video-based asynchronous neurological examination (VANE) was also administered.^18^ Functional and behavioral scales included the Geriatric Depression Scale (GDS),^19^ complemented by study partner questionnaires that included the neuropsychiatric inventory (NPI)^20^ and the functional assessment questionnaire (FAQ).^21^ We complemented these NACC-UDS informant questionnaires with the Quick Dementia Rating System (QDRS).^22^

Cognitive outcomes were defined using the 2011 National Institute of Aging/Alzheimer’s Association recommendations for mild cognitive impairment (MCI)^23^ and NACC-UDS battery for dementia^24^. Cognitive impairment syndromes were defined as follows using the NACC-UDS forms nomenclature: No cognitive impairment (NoCI), defined by global clinical dementia rating (CDR) = 0 and normal or mostly normal cognitive testing; MCI, defined by the presence of cognitive concerns or objective impairment on testing when impairments were consistently present within a given domain (memory, language, visuospatial, attention, and/or executive), but preserved independence of functional abilities (CDR = 0.5); and dementia, defined by global CDR = 1 or higher (significant functional impairment) and impairment in one or more cognitive domains. MCI was further classified as non-amnestic MCI or amnestic MCI (**Supplemental Table 2**). When participants did not meet any of the above categories they were classified as having cognitive impairment (CI)–not MCI (CI–noMCI). The diagnostic categories were assigned through consensus. A consensus committee of five adjudicators, all of whom have extensive experience with UDS-based assessments, reviewed all neuropsychological and functional measures. The adjudicators were masked to DPP treatment assignment, metformin treatment, and diabetes status. Cognitive impairment syndromes were further classified as amyloid positive or negative using plasma p-tau 217 measured at the time of cognitive assessments. We selected p-tau217 based on its analytical performance, including longitudinal assay precision (coefficient of variation, 5.03%), using the Lumipulse platform (Fujirebio, Malvern, PA, USA). We used a cutoff of 0.23 pg/mL, corresponding to previously validated thresholds to define amyloid positivity.^25^

The secondary cognitive outcomes were based on tests of executive function and memory (i.e., the Digit Symbol Substitution Test^26^ [DSST] and the Spanish English Verbal Learning Test^27^ [SEVLT] respectively), which were administered five times (approximately every two years) between 2009 and 2024. The DSST was used to ascertain executive performance, and the sum of words recalled across the three immediate recall trials on the SEVLT (SEVLT-IR) was used to ascertain memory performance. Both were reported as continuous measures, with higher scores indicating better performance (DSST range, 0–93; SEVLT-IR range, 0–45).

### Covariates

We included covariates to assess balance and differential attrition across randomization groups and provide conditional effect estimates. Baseline demographic and clinical covariates were compared across randomization groups including age, sex race/ethnicity, education, income category, smoking history, body mass index (BMI), LDL cholesterol, SF-36 Health Survey Mental Component Summary score, and APOE-ε4 carrier status. Age at each assessment was compared across randomization groups.

### Statistical analyses

We compared characteristics at the time of randomization and at the 2022–2024 DPPOS study visit among the placebo, metformin, and ILS groups using analysis of covariance for continuous variables and chi-squared tests for categorical variables. Our primary analyses examined associations of randomization to metformin vs. placebo and ILS with cognitive syndromes by estimating odds ratios (OR) with multinomial logistic regression using NoCI as the reference category because the assumption of proportional odds did not hold. In secondary analyses for cognitive performance, we examined associations of randomization to metformin vs. placebo and ILS with changes in the DSST and SEVLT-IR scores using linear mixed-effects models with a compound symmetry covariance structure to account for the longitudinal measures. We reported the coefficients for metformin, time, and interaction terms of metformin with time. A statistically significant term for metformin indicated significant differences in DSST or SEVLT-IR scores compared with the comparison group, with positive coefficients indicating higher (better) scores in the metformin group. A statistically significant term for time indicated significant cognitive performance changes over time; improvement in scores if positive, worsening in scores if negative. A statistically significant interaction term indicated significant differences in the slope of cognitive change between the metformin and the placebo and/or ILS groups. We used R version 4.2.1 with p<0.05 considered statistically significant, without adjustment for multiple comparisons.

## RESULTS

At randomization, the median age of the 1,483 DPP/DPPOS participants with cognitive testing results was 48 (IQR 42, 54) years. Most (74%) were women and 51% were White, 21% Black, and 17% Hispanic/Latino. Their median BMI was 33 kg/m² and 24% carried the APOE-ε4 variant. There were no significant differences in age, sex, race/ethnicity, BMI, LDL-cholesterol, income, education, smoking prevalence, or APOE-ε4 carrier frequency across treatment groups. In 2009–2010, when the DSST and SEVLT were first measured, the median age was 60 (IQR 54, 66) years with no significant differences across treatment groups; 48% had diabetes (58%, 49%, and 47% in placebo, metformin, and ILS groups respectively, p=0.01). At the 2022–2024 visits, when cognitive outcomes were measured and adjudicated, the median age was 74 (IQR 68, 79) years, again with no significant differences across treatment groups. Diabetes had developed in 72% of the cohort (75%, 68%, and 71% in placebo, metformin, and ILS groups, respectively, p=0.03, **Table 1**).

**Table 1.**
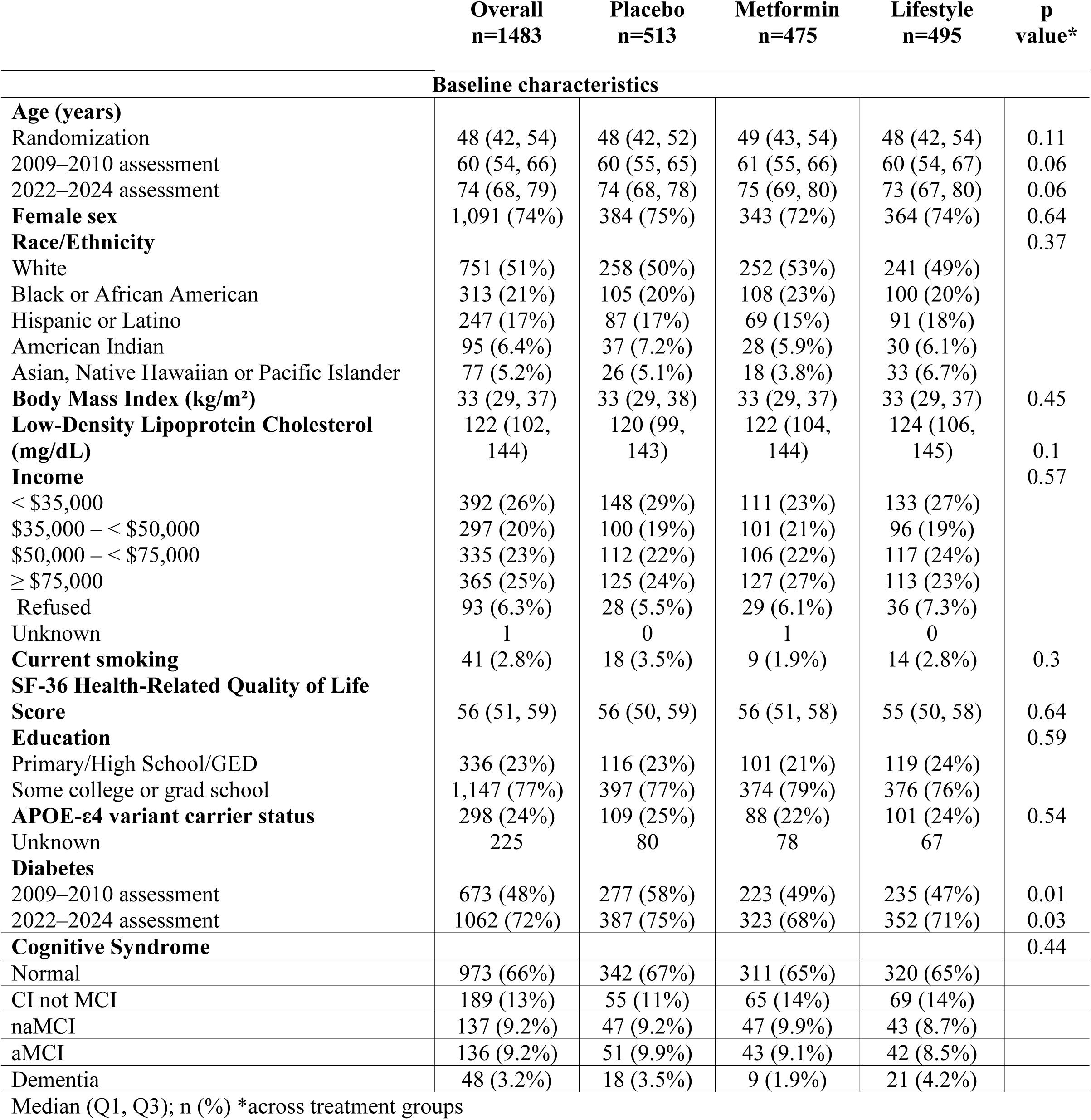
Characteristics of DPPOS participants, overall and stratified by original DPP treatment assignment.

The distribution of cognitive impairment syndromes assessed in 2022–2024 did not differ significantly by treatment group (**Fig. 1**); however, randomization to metformin was associated with a 60% (OR 0.40 [95% confidence interval 0.17, 0.97]) and 62% (OR 0.38 [95% confidence interval 0.16, 0.89]) lower odds of dementia compared with placebo and ILS, respectively, after adjustment for demographics, education, income, and APOE-ε4 genotype (**Table 2**). As an exploratory analysis, counts and proportions of cognitive impairment syndrome categories stratified by plasma p-tau217 concentration were summarized across treatment groups. Among participants with plasma p-tau 217 measured at the time of cognitive assessment amyloid-negative dementia was observed in 3 (1.4%), 2 (0.8%), and 3 (1.3%) participants in the metformin, placebo, and ILS groups respectively, whereas amyloid-positive dementia was observed in 3 (1.4%), 10 (4.0%), and 12 (4.9%) respectively.

**Fig. 1.**
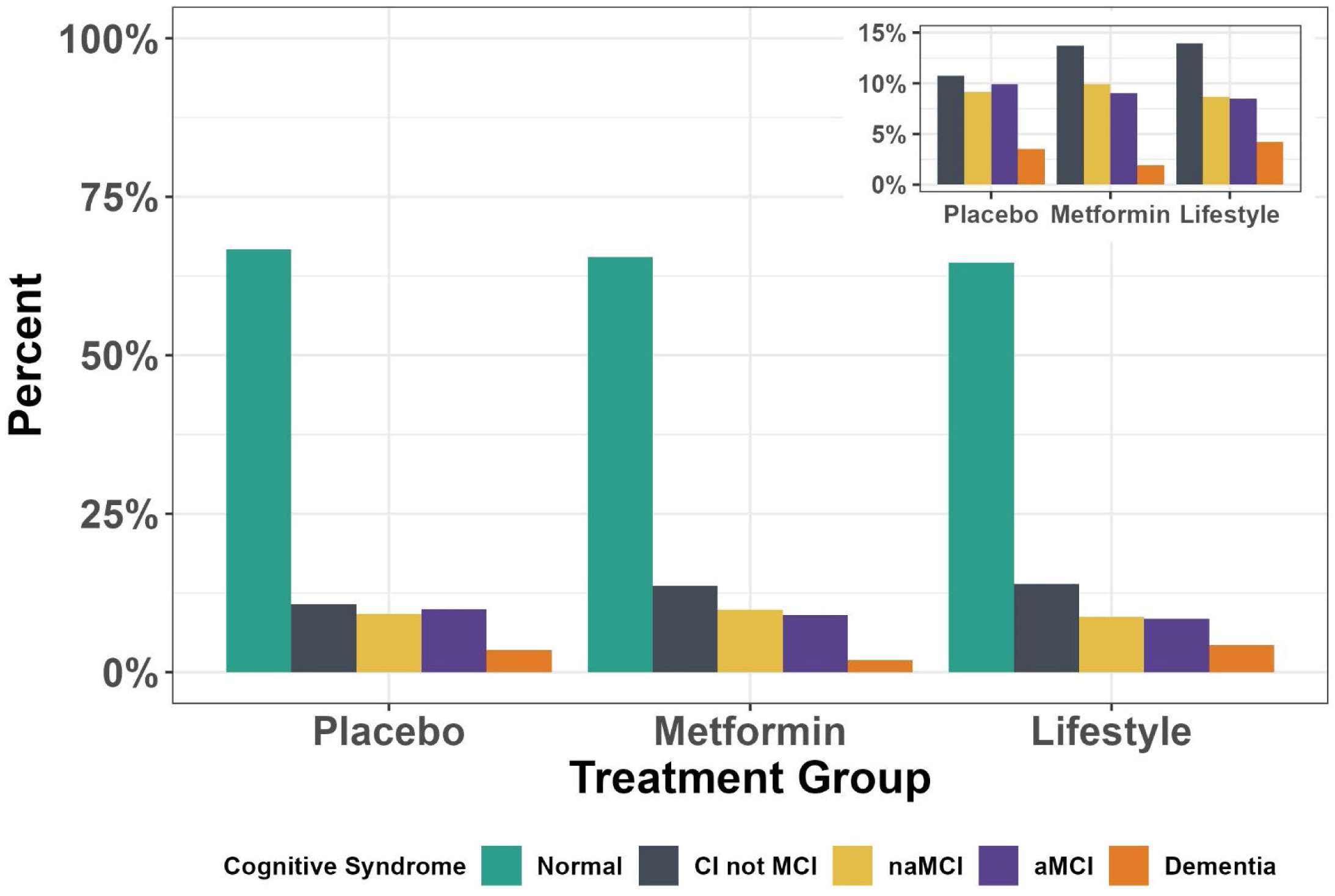
Prevalence of Cognitive Impairment Syndromes by DPP Randomized Treatment Group (will add gradients for journal submission) Distribution of cognitive impairment syndromes at the DPPOS 2022–2024 visit stratified by treatment assignment. Inset panel excludes individuals in the no cognitive impairment category. The frequency distributions of the cognitive syndromes did not differ significantly by treatment group, either unadjusted (p=0.45), adjusted for age, sex, race/ethnicity, education, and income (p=0.34), or also adjusted for APOE-Ɛ4 carrier status (p=0.31). However, the associations of dementia with metformin compared with either placebo or ILS were significant (95% confidence intervals for odds ratios excluded 1, uncorrected for multiple comparisons) in adjusted models, as shown in Table 2.

**Table 2.** OR and 95% confidence interval relating DPP randomization to metformin with cognitive impairment syndromes at the 2022–2024 visit in DPPOS.

| Outcome | Metformin vs Placebo |  |  | Metformin vs Lifestyle |  |  |
| --- | --- | --- | --- | --- | --- | --- |
|  | Model 1 | Model 2 | Model 3 | Model 1 | Model 2 | Model 3 |
| CI not MCI vs Normal | 1.25(0.85, 1.85) | 1.25 (0.84, 1.87) | 1.25 (0.84, 1.87) | 0.95 (0.65, 1.38) | 0.95 (0.65, 1.39) | 0.95 (0.64, 1.39) |
| naMCI vs Normal | 1.09 (0.71, 1.68) | 1.05 (0.67, 1.65) | 1.04 (0.66, 1.62) | 1.15(0.74, 1.79) | 1.11 (0.70, 1.77) | 1.11 (0.70, 1.76) |
| aMCI vs Normal | 0.92(0.60, 1.42) | 0.82 (0.52, 1.29) | 0.80 (0.51, 1.27) | 1.03 (0.65, 1.61) | 0.95 (0.60, 1.53) | 0.94 (0.58, 1.51) |
| Dementia vs Normal | 0.55 (0.24, 1.23) | **0.41 (0.18, 0.98)** | **0.40 (0.17, 0.97)** | **0.44 (0.20, 0.97)** | **0.40 (0.17, 0.92)** | **0.38 (0.16, 0.89)** |
| p-value* | 0.45 | 0.34 | 0.31 | 0.45 | 0.34 | 0.31 |

Model 1: Unadjusted
Model 2: age + sex + race/ethnicity + education + income category
Model 3: Model 2 + APOE-ε4 genotype
Abbreviations: CL (confidence limits), OR (odds ratio), Cognitive impairment not mild cognitive impairment (CI not MCI) aMCI (amnesic MCI), naMCI (non-amnesic MCI), normal (no cognitive impairment).
The sample sizes of the five cognitive categories are: Normal: n=973; CI not MCI: n=189; naMCI: n=137; aMCI: n=136; Dementia: n=48
\* Omnibus p-value from likelihood ratio test

The longitudinal trajectories in executive function from the DSST and memory from the SEVLT-IR showed declines in all three groups with parallel mean trajectories (all p values for interaction> 0.05) (**Fig. 2**). However, longitudinal performance in the SEVLT-IR was significantly better in the metformin group compared with the placebo group (β=0.58; 95% confidence interval: 0.09, 1.1; p=0.02, **Table 3**), although this difference was modest (Cohen’s d=0.1). There was no evidence of differences between ILS and placebo across analyses (**Supplemental Tables 3 and 4**).

**Fig. 2.**
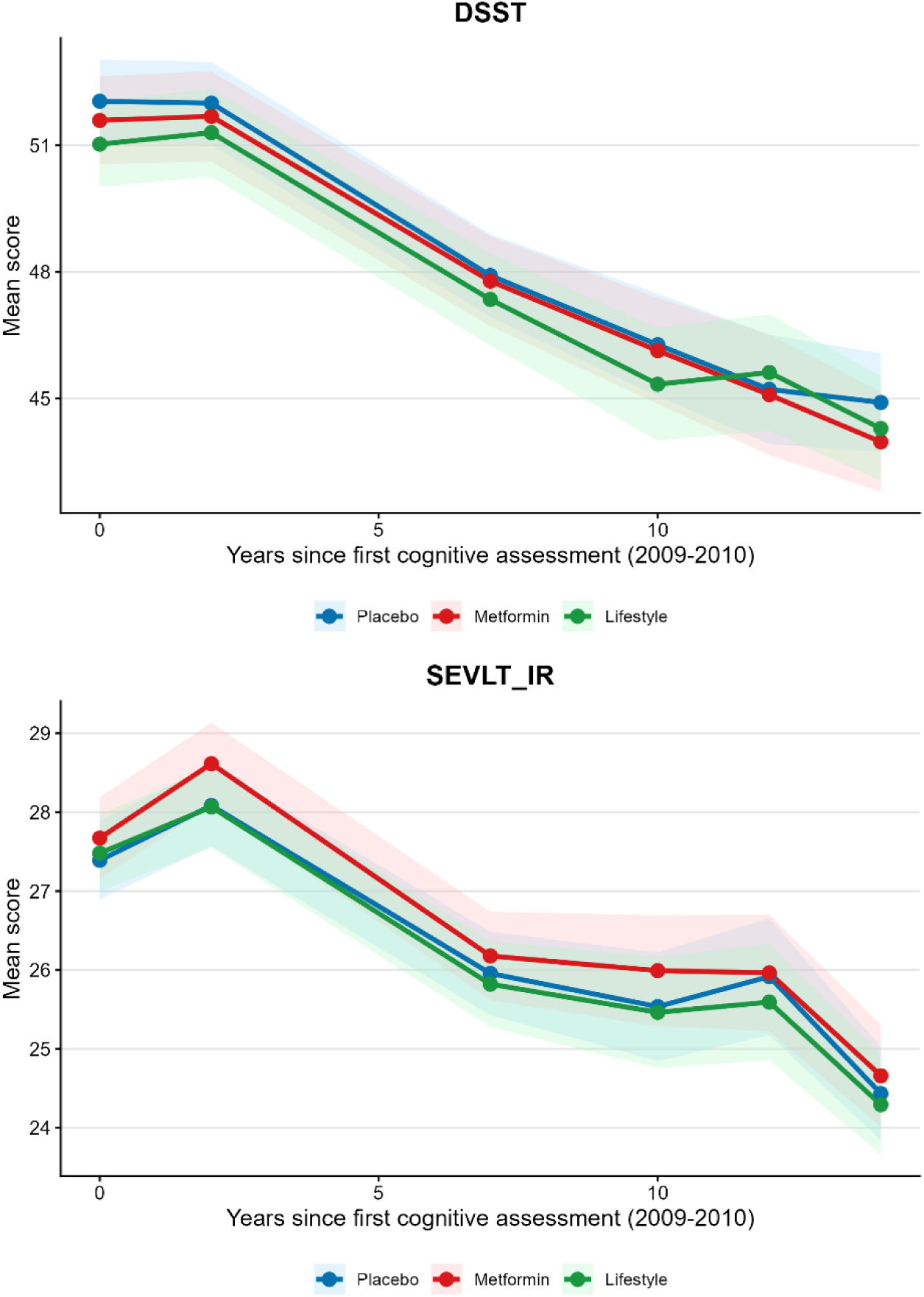
Mean DSST and SEVLT-IR scores with 95% confidence intervals over time by DPP randomized treatment group. Longitudinal trajectories in mean (A) Digit Symbol Substitution Test and (B) Spanish English Verbal Learning Test—Immediate Recall scores among DPPOS participants (2009–2024), stratified by treatment assignment.

**Table 3.** Estimates and 95% confidence interval relating original DPP treatment assignment with amnestic and non-amnestic cognitive performance from 2009–2024 in participants with cognitive outcomes in 2022–2024.

| Outcome |  | n* | Metformin vs Placebo |  |  | Metformin vs Lifestyle |  |  |
| --- | --- | --- | --- | --- | --- | --- | --- | --- |
|  |  |  | estimate | 95% CL | p value | estimate | 95% CL | p value |
| Digit Symbol Substitution Test Score | Model 1 | 1654 | -0.24 | (-1.6, 1.1) | 0.7 | 0.61 | (-0.73, 2.0) | 0.4 |
|  | Model 2 | 1653 | 0.28 | (-0.76, 1.3) | 0.6 | 0.66 | (-0.39, 1.7) | 0.2 |
|  | Model 3 | 1653 | 0.27 | (-0.76, 1.3) | 0.6 | 0.65 | (-0.40, 1.7) | 0.2 |
| Spanish English Verbal Learning Test — Immediate Recall Score | Model 1 | 1668 | 0.41 | (-0.22, 1.0) | 0.2 | 0.43 | (-0.20, 1.10) | 0.2 |
|  | Model 2 | 1667 | 0.58 | (0.09, 1.1) | **0.02** | 0.44 | (-0.06, 0.94) | 0.08 |
|  | Model 3 | 1667 | 0.58 | (0.09, 1.1) | **0.02** | 0.44 | (-0.06, 0.94) | 0.08 |

\*corresponds to approximately 4.8 measures per participant
Model 1: visit year + treatment assignment
Model 2: Model 1+ age + sex + race + education + income category
Model 3: Model 2 + APOE ε4 carrier status

## DISCUSSION

To our knowledge, this is the first report of the association of metformin with cognitive impairment syndromes, including dementia, in the context of long-term follow-up of a randomized controlled trial using state-of-the-art methods to ascertain cognitive syndromes. Although the overall distributions of the cognitive syndromes did not differ significantly by treatment group, randomization to metformin, resulting in an average 15 years/person of metformin use, was associated with an approximately 60% lower odds of dementia compared with placebo and ILS. Randomization to metformin was also associated with higher longitudinal mean scores on a measure of memory performance preceding and including the time when cognitive syndromes were ascertained. Although the magnitude of the difference was small, it was comparable to the effect found in recent clinical trials of multimodal interventions for the prevention of cognitive decline.^28,29^ Importantly, we found no evidence of harmful effects of metformin on cognition.

Observational studies have provided conflicting data on the risk of dementia related to metformin^8^ with some reporting an increased^9,10^ and some reporting decreased^11^ risk. Most observational studies have been limited by lack of control for confounding by indication, use of administrative data for the ascertainment of metformin exposure and/or dementia, or both issues.^8^ One study using administrative data reported that metformin cessation among persons with T2D was associated with a higher risk of dementia.^30^ Studies in persons with diabetes that have attempted to account for confounding by indication (e.g., propensity score methods) have reported a lower risk of dementia in relation to metformin^11^ but are still limited by use of administrative data. Our study overcomes these limitations by examining randomized metformin to ascertain metformin exposure and using state-of-the-art cognitive assessments to ascertain cognitive outcomes. We previously reported that randomization to metformin was not associated with cognitive performance in the SEVLT and DSST in 2009–2010 when participants’ mean age was 60 years,^31^ with a non-significant separation favoring metformin for the SEVLT-IR in 2011– 2012.^31^ The difference in SEVLT-IR has become significant in our current analyses that include approximately five waves of SEVLT data over approximately 10 years of follow-up.

There is a dearth of randomized clinical trial data on the effects of metformin on cognitive outcomes. A small 12-month 1:1 placebo-controlled randomized trial of metformin in 80 participants with amnestic MCI and without diabetes reported better performance at 12 months in the Buchske Selective Reminding Test,^13^ a test similar to the SEVLT used in DPPOS.

Two observational studies^32,33^ have reported that diabetes treatment (most often with metformin) is associated with lower brain amyloid burden compared with untreated diabetes. Animal studies conflict on whether metformin increases or reduces amyloid burden in the brain, the main culprit of AD. ^34^ Although the number of dementia cases in each group in DPPOS was small, the exploratory descriptive analysis stratified by p-tau217 positivity suggests that the difference in dementia by treatment group was driven by a relatively lower proportion of AD (amyloid-positive) related dementia cases among participants originally randomized to metformin.

Several issues require further consideration. Most importantly, the overall prevalences of cognitive impairment syndromes did not differ by treatment group, although randomization to metformin was associated with lower odds of dementia compared with placebo and ILS, but only in analyses uncorrected for multiple comparisons. Stringent adjustments for multiple comparisons increase type 2 error and may miss the opportunity to detect a real association or phenomenon. Hence, some have recommended not adjusting for multiple comparisons.^35^ We would contend that the concordant findings from the primary and secondary analyses reported herein suggest that metformin decreases the risk of dementia. We did not find an association of metformin with the risk of amnestic MCI, which is a prodromal phase for those at high risk of progressing to dementia.^23^ It is possible that metformin has a threshold effect that lowers the risk of progression from amnestic MCI to dementia but does not slow the progression from NoCI to amnestic MCI. We cannot examine this hypothesis with a single wave of cognitive syndrome adjudication and require longer follow-up to enable these analyses.

This study has several strengths. The exposure was the randomized DPP treatment assignment, which reduced the potential for confounding by indication, a major limitation of existing evidence in this area. Compared to previous retrospective studies of metformin and cognition, which relied largely on claims data to quantify metformin exposure, metformin use was assessed with structured pill counts during DPP and DPPOS and quantified by calculating cumulative person years of treatment for both study and out-of-study use. NACC-UDS definitions, operationalized with standardized clinical criteria implemented across Alzheimer’s Disease Research Centers in the United States, were used to assess cognitive outcomes.

Employing standardized neuropsychological measures with central adjudication provided direct assessment of cognitive performance and outcomes, reducing detection and misclassification biases inherent in claims-based proxies for cognitive outcomes. Finally, detailed phenotyping of the DPP/DPPOS cohort over decades allowed some assessment of differential loss to follow-up by randomized treatment group.

This study also has limitations. An important limitation is that we analyzed long-term follow-up data, which is subject to common problems in both randomized and observational settings, including departures from assigned treatments due to changing medical conditions and loss of participants due to death or withdrawal.^15^ Many participants not originally assigned to metformin “crossed over” to metformin therapy, usually after developing diabetes, which potentially weakened causal inference. Although out-of-study metformin use could dilute the true effects of randomized metformin, metformin exposure remained substantially greater in the metformin group than in either the placebo or ILS group. Cognitive outcomes were not collected at baseline or early follow-ups. If such outcomes impacted continued participation and occurred differentially by treatment group early in the study, this would introduce survivorship bias by altering who remained under observation. The possibility of residual confounding cannot be excluded; however, baseline characteristics among the population that remained in the analysis at the 2022–2024 visit were well balanced across treatment groups. Finally, rates of abnormal cognitive outcomes were low, limiting power to detect differences across treatment groups.

Despite these limitations, our data suggest that this long-term follow-up of a randomized clinical trial provides the best available evidence for metformin effects on dementia.

In conclusion, in this multicenter randomized clinical trial and its long-term follow-up study, randomization to metformin, representing approximately 15 years of average metformin exposure, was associated with lower odds of dementia ascertained at 22–24 years of follow-up and with higher mean scores on a measure of memory performance over time. Together, these findings suggest a potential long-term protective effect of metformin on cognition among adults at high risk for T2D at baseline. Longer follow-up with more dementia cases is needed to confirm our findings.

## Supporting information

DPPOS Investigators Appendix

## ACKNOWLEDGMENTS

The DPP Research Group gratefully acknowledges the commitment and dedication of the participants of the DPP and DPPOS.

## Author Contributions

Concept and design: Knowler, Luchsinger, Nathan, Wander

Acquisition, analysis, or interpretation of data: Wander, Doherty, Pan, Carmichael, Kuo, Munshi, Wallia, Noble, Shah, Nadkarni, Mudaliar, Dabelea, Temprosa, Nathan, Luchsinger

Drafting of the manuscript: Knowler, Luchsinger, Nathan, Wander

Critical revision of the manuscript for important intellectual content: Wander, Doherty, Pan, Carmichael, Turner, Kuo, Munshi, Wallia, Noble, Shah, Nadkarni, Mudaliar, Dabelea, Temprosa, Knowler, Nathan, Luchsinger

Statistical analysis: Doherty, Pan, Temprosa

Obtained funding: Luchsinger, Nathan, Temprosa

Administrative, technical, or material support: Luchsinger, Nathan, Temprosa

Supervision: Luchsinger, Nathan

## Additional Information

co-author William C. Knowler passed away on July 7^th^, 2026, when the last draft of the manuscript had been reviewed before submission.

## STATEMENT OF ETHICS

The study was conducted according to the guidelines set out in the Declaration of Helsinki and all procedures involving research study participants. Prior to initiating the study protocol, each participant provided written informed consent and each study center obtained approval from its respective institutional review board. The trials are registered at ClinicalTrials.gov (Diabetes Prevention Program: NCT00004992; Diabetes Prevention Program Outcomes Study: NCT00038727; Diabetes Prevention Program Outcomes Study AD/ADRD: NCT05704309).

## DATA AVAILABILITY STATEMENT

In accordance with the NIH Public Access Policy, we continue to provide all manuscripts to PubMed Central including this manuscript DPP/DPPOS has provided the protocols and lifestyle and medication intervention manuals to the public through its public website (https://www.dppos.org). The DPPOS abides by the NIDDK data sharing policy and implementation guidance as required by the NIH/NIDDK (https://repository.niddk.nih.gov/studies/dppos/)

## REFERENCES

1. 9. Pharmacologic Approaches to Glycemic Treatment: Standards of Care in Diabetes-2026. Diabetes Care. Jan 1 2026;49(Supplement_1):S183–s215. doi:10.2337/dc26-S009

2. Xing C, Zhao H, Zhang J, He B. Effect of metformin versus metformin plus liraglutide on gonadal and metabolic profiles in overweight patients with polycystic ovary syndrome. Front Endocrinol (Lausanne). 2022;13:945609. doi:10.3389/fendo.2022.945609

3. Knowler WC, Barrett-Connor E, Fowler SE, et al. Reduction in the incidence of type 2 diabetes with lifestyle intervention or metformin. N Engl J Med. Feb 7 2002;346(6):393–403. doi:10.1056/NEJMoa012512

4. Crandall JP, Dabelea D, Knowler WC, Nathan DM, Temprosa M, Group DPPR. The Diabetes Prevention Program and Its Outcomes Study: NIDDK’s Journey Into the Prevention of Type 2 Diabetes and Its Public Health Impact. Diabetes Care. 2025;48(7):1101–1111. doi:10.2337/dc25-0014

5. Knowler WC, Fowler SE, Hamman RF, et al. 10-year follow-up of diabetes incidence and weight loss in the Diabetes Prevention Program Outcomes Study. Lancet. Nov 14 2009;374(9702):1677–86. doi:10.1016/s0140-6736(09)61457-4

6. Knowler WC, Doherty L, Edelstein SL, et al. Long-term effects and effect heterogeneity of lifestyle and metformin interventions on type 2 diabetes incidence over 21 years in the US Diabetes Prevention Program randomised clinical trial. The Lancet Diabetes & Endocrinology. 2025;13(6):469–481. doi:10.1016/S2213-8587(25)00022-1

7. Lv Z, Guo Y. Metformin and Its Benefits for Various Diseases. Front Endocrinol (Lausanne). 2020;11:191. doi:10.3389/fendo.2020.00191

8. Tahmi M, Benitez R, Luchsinger JA. Metformin as a Potential Prevention Strategy for Alzheimer’s Disease and Alzheimer’s Disease Related Dementias. J Alzheimers Dis. 2024;101(s1):S345–s356. doi:10.3233/jad-240495

9. Imfeld P, Bodmer M, Jick SS, Meier CR. Metformin, other antidiabetic drugs, and risk of Alzheimer’s disease: a population-based case-control study. Research Support, Non-U.S. Gov’t. Journal of the American Geriatrics Society. May 2012;60(5):916–21. doi:10.1111/j.1532-5415.2012.03916.x

10. Moore EM, Mander AG, Ames D, et al. Increased Risk of Cognitive Impairment in Patients With Diabetes Is Associated With Metformin. Diabetes Care. 2013;36(10):2981–2987.

11. Orkaby AR, Cho K, Cormack J, Gagnon DR, Driver JA. Metformin vs sulfonylurea use and risk of dementia in US veterans aged ≥65 years with diabetes. Neurology. 2017;89(18):1877. doi:10.1212/WNL.0000000000004586

12. Zhang Y, Zhang Y, Shi X, et al. Metformin and the risk of neurodegenerative diseases in patients with diabetes: A meta-analysis of population-based cohort studies. Diabet Med. Jun 2022;39(6):e14821. doi:10.1111/dme.14821

13. Luchsinger JA, Perez T, Chang H, et al. Metformin in Amnestic Mild Cognitive Impairment: Results of a Pilot Randomized Placebo Controlled Clinical Trial. J Alzheimers Dis. 2016;51(2):501–14. doi:10.3233/jad-150493

14. Koenig AM, Mechanic-Hamilton D, Xie SX, et al. Effects of the Insulin Sensitizer Metformin in Alzheimer Disease: Pilot Data From a Randomized Placebo-controlled Crossover Study. Alzheimer Dis Assoc Disord. Apr–Jun 2017;31(2):107–113. doi:10.1097/WAD.0000000000000202

15. Knowler WC, Pan Q, Shu S, et al. Analysis of Long-term Follow-up of a Randomized Clinical Trial With Departures From Assigned Treatments: Estimation of Metformin Effects on Diabetes and Its Complications in the Diabetes Prevention Program Outcomes Study. Diabetes Care. Oct 1 2025;48(10):1668–1675. doi:10.2337/dci25-0032

16. Weintraub S, Besser L, Dodge HH, et al. Version 3 of the Alzheimer Disease Centers’ Neuropsychological Test Battery in the Uniform Data Set (UDS). Alzheimer Dis Assoc Disord. 2018;32(1):10–17. doi:10.1097/WAD.0000000000000223

17. Doherty L, Dechiario I, Sherif H, et al. Implementing the National Alzheimers Coordinating Center Uniform Data Set (v3) within the Diabetes Prevention Program Outcomes Study. medRxiv. 2026:2026.07.17.26357765. doi:10.64898/2026.07.17.26357765

18. Noble JM, Nadkarni NK, Martinez D, et al. Implementation of a standardized Video-based Asynchronous Neurological Examination (VANE) in a multi-center observational study of Alzheimers disease (AD) and AD related dementias. medRxiv. 2026:2026.07.15.26357456. doi:10.64898/2026.07.15.26357456

19. Yesavage JA, Brink TL, Rose TL, et al. Development and validation of a geriatric depression screening scale: a preliminary report. J Psychiatr Res. 1982;17(1):37–49.

20. Cummings JL. The Neuropsychiatric Inventory: assessing psychopathology in dementia patients. Neurology. May 1997;48(5 Suppl 6):S10–6. doi:10.1212/wnl.48.5_suppl_6.10s

21. Pfeffer RI, Kurosaki TT, Harrah CH, Jr., Chance JM, Filos S. Measurement of functional activities in older adults in the community. J Gerontol. May 1982;37(3):323–9.

22. Galvin JE. THE QUICK DEMENTIA RATING SYSTEM (QDRS): A RAPID DEMENTIA STAGING TOOL. Alzheimers Dement (Amst). 2015;1(2):249–259. doi:10.1016/j.dadm.2015.03.003

23. Albert MS, DeKosky ST, Dickson D, et al. The diagnosis of mild cognitive impairment due to Alzheimer’s disease: recommendations from the National Institute on Aging-Alzheimer’s Association workgroups on diagnostic guidelines for Alzheimer’s disease. Alzheimers Dement. May 2011;7(3):270–9. doi:10.1016/j.jalz.2011.03.008

24. McKhann GM, Knopman DS, Chertkow H, et al. The diagnosis of dementia due to Alzheimer’s disease: recommendations from the National Institute on Aging-Alzheimer’s Association workgroups on diagnostic guidelines for Alzheimer’s disease. Alzheimers Dement. May 2011;7(3):263–9. doi:10.1016/j.jalz.2011.03.005

25. Feizpour A, Doecke JD, Doré V, et al. Detection and staging of Alzheimer’s disease by plasma pTau217 on a high throughput immunoassay platform. EBioMedicine. Nov 2024;109:105405. doi:10.1016/j.ebiom.2024.105405

26. Wechsler D. Wechsler Adult Intelligence Scale-Revised. Psychological Corporation; 1988.

27. Gonzalez HM, Mungas D, Reed BR, Marshall S, Haan MN. A new verbal learning and memory test for English- and Spanish-speaking older people. J Int Neuropsychol Soc. Jul 2001;7(5):544–55.

28. Ngandu T, Lehtisalo J, Solomon A, et al. A 2 year multidomain intervention of diet, exercise, cognitive training, and vascular risk monitoring versus control to prevent cognitive decline in at-risk elderly people (FINGER): a randomised controlled trial. The Lancet. 2015;385(9984):2255–2263. doi:10.1016/S0140-6736(15)60461-5

29. Baker LD, Espeland MA, Whitmer RA, et al. Structured vs Self-Guided Multidomain Lifestyle Interventions for Global Cognitive Function: The US POINTER Randomized Clinical Trial. JAMA. 2025;334(8):681–691. doi:10.1001/jama.2025.12923

30. Zimmerman SC, Ferguson EL, Choudhary V, et al. Metformin Cessation and Dementia Incidence. JAMA Network Open. 2023;6(10):e2339723–e2339723. doi:10.1001/jamanetworkopen.2023.39723

31. Luchsinger JA, Ma Y, Christophi CA, et al. Metformin, Lifestyle Intervention, and Cognition in the Diabetes Prevention Program Outcomes Study. 10.2337/dc16-2376. Diabetes Care. 2017;

32. Akinci M, Aziz F, Guzman D, et al. Association of type 2 diabetes treatment status with in vivo biomarkers of Alzheimer’s disease. Alzheimers Dement. Feb 2026;22(2):e71214. doi:10.1002/alz.71214

33. McIntosh EC, Nation DA, for the Alzheimer’s Disease Neuroimaging I. Importance of Treatment Status in Links Between Type 2 Diabetes and Alzheimer’s Disease. Diabetes Care. 2019;42(5):972–979. doi:10.2337/dc18-1399

34. Jack CR, Jr., Bennett DA, Blennow K, et al. NIA-AA Research Framework: Toward a biological definition of Alzheimer’s disease. Alzheimers Dement. Apr 2018;14(4):535–562. doi:10.1016/j.jalz.2018.02.018

35. Rothman KJ. No adjustments are needed for multiple comparisons. Epidemiology. Jan 1990;1(1):43–6.

