## Supplemental Tables and Figures for "Randomized metformin and cognitive outcomes in the Diabetes Prevention Program Outcomes Study"

**Supplemental Table 1**. Comparison of baseline characteristics of DPP participants included in the analysis with those not included

| **Participant Characteristic** | **Overall** N = 3,234*^1^* | **Included in Analysis** N = 1,483*^1^* | **Not Included** N = 1,751*^1^* | **p-value***^2^* |
| --- | --- | --- | --- | --- |
| **Age at Randomization** | 50 (43, 58) | 48 (42, 54) | 52 (44, 62) | **<0.001** |
| **Treatment Group** |  |  |  | 0.34 |
| Placebo | 1,082 (33%) | 513 (35%) | 569 (32%) |  |
| Metformin | 1,073 (33%) | 475 (32%) | 598 (34%) |  |
| Lifestyle | 1,079 (33%) | 495 (33%) | 584 (33%) |  |
| **Sex** |  |  |  | **<0.001** |
| Male | 1,043 (32%) | 392 (26%) | 651 (37%) |  |
| Female | 2,191 (68%) | 1,091 (74%) | 1,100 (63%) |  |
| **Race/Ethnicity** |  |  |  | **<0.001** |
| White | 1,768 (55%) | 751 (51%) | 1,017 (58%) |  |
| Black or African American | 645 (20%) | 313 (21%) | 332 (19%) |  |
| Hispanic or Latino | 508 (16%) | 247 (17%) | 261 (15%) |  |
| American Indian | 171 (5.3%) | 95 (6.4%) | 76 (4.3%) |  |
| Asian, Native Hawaiian or Pacific Islander | 142 (4.4%) | 77 (5.2%) | 65 (3.7%) |  |
| **BMI** | 33 (29, 37) | 33 (29, 37) | 33 (29, 37) | 0.83 |
| **Low-Density Lipoprotein Cholesterol (mg/dL)** | 123 (102,145) | 122 (102,144) | 124 (102,146) | 0.24 |
| **Income** |  |  |  | **<0.001** |
| <$35,000 | 1,007 (31%) | 392 (26%) | 615 (35%) |  |
| $35,000 - <50,000 | 641 (20%) | 297 (20%) | 344 (20%) |  |
| $50,000 - <75,000 | 646 (20%) | 335 (23%) | 311 (18%) |  |
| $75,000+ | 682 (21%) | 365 (25%) | 317 (18%) |  |
| Refused | 257 (7.9%) | 93 (6.3%) | 164 (9.4%) |  |
| Unknown | 1 | 1 | 0 |  |
| **Smoking** | 226 (7.0%) | 84 (5.7%) | 142 (8.1%) | **0.007** |
| **SF-36 Health-Related Quality of Life Score** | 56 (51,59) | 56 (51,59) | 56 (52,59) |  |
| **Education** |  |  |  | **<0.001** |
| Primary/High School/GED | 834 (26%) | 336 (23%) | 498 (28%) |  |
| Some college or grad school | 2,400 (74%) | 1,147 (77%) | 1,253 (72%) |  |
| **APOE ε4 variant** | 721 (27%) | 298 (24%) | 423 (29%) | **0.001** |
| Unknown | 522 | 225 | 297 |  |
| *^1^*Median (Q1, Q3); n (%) | | | | |
| ^2^Wilcoxon rank sum test; Pearson's Chi-squared test  Of those not included, 598 had died by 2022. | | | | |

| **Supplemental Table 2.** Cognitive impairment syndrome definitions | |
| --- | --- |
| **Construct** | **Definitions or Measures** |
| Cognitive impairment syndrome categories | - Normal cognitive impairment (normal) |
|  | - CI not mild cognitive impairment (MCI) |
|  | - MCI |
|  | ·  Non-amnestic MCI (naMCI) |
|  | ·  Amnestic MCI (aMCI) |
|  | - Dementia |
| **Exploratory** Cognitive impairment syndrome categories split by amyloid positivity/negativity | - Normal cognition / Amyloid - (ref) |
|  | - Normal cognition / Amyloid + |
|  | - CI not MCI / Amyloid - |
|  | - CI not MCI / Amyloid + |
|  | - naMCI / Amyloid - |
|  | - naMCI / Amyloid + |
|  | - aMCI / Amyloid - |
|  | - aMCI / Amyloid + |
|  | - Dementia / Amyloid - |
|  | - Dementia / Amyloid + |
| Cognitive performance | - Amnestic Cognitive Performance: Longitudinal SEVLT (first 3 trials-immediate recall) 2009–2024 |
|  | - Non-amnestic Cognitive Performance: Longitudinal DSST 2009–2024 |

| **Supplemental Table 3.** OR and 95% confidence interval relating randomization to lifestyle vs. placebo with cognitive impairment syndromes at the 2022–2024 visit in DPPOS | | | |
| --- | --- | --- | --- |
|  | Lifestyle vs Placebo | | |
| Outcome | Model 1 | Model 2 | Model 3 |
| CI not MCI vs Normal | 1.31 (0.85, 1.85) | 1.32 (0.89, 1.96) | 1.32 (0.89, 1.96) |
| naMCI vs Normal | 0.95 (0.61, 1.48) | 0.95 (0.60, 1.49) | 0.93 (0.59, 1.48) |
| aMCI vs Normal | 0.90 (0.58, 1.39) | 0.86 (0.54, 1.35) | 0.86 (0.54, 1.35) |
| Dementia vs Normal | 1.24 (0.65, 2.38) | 1.05 (0.53, 2.09) | 1.07 (0.53, 2.18) |
| p-value* | 0.45 | 0.34 | 0.31 |
| Model 1: Unadjusted | | | |
| Model 2: age + sex + race/ethnicity + education + income category | | | |
| Model 3: Model 2 + APOE-Ɛ4 genotype | | | |
| Abbreviations: CL (confidence limits), OR (odds ratio), Cognitive impairment not mild cognitive impairment (CI not MCI) aMCI (amnestic MCI), naMCI (non-amnestic MCI), normal (no cognitive impairment).  The sample sizes of the five cognitive categories are: Normal: n=973; CI not MCI: n=189; naMCI: n=136; aMCI: n=137; Dementia: n=48 | | | |
| * Omnibus p-value from likelihood ratio test | | | |

| **Supplemental Table 4.** Estimates and 95% confidence interval relating original DPP treatment assignment to lifestyle vs. placebo with amnestic and non-amnestic cognitive performance from 2009–2024 in participants with cognitive outcomes in 2022–2024 | | | | |
| --- | --- | --- | --- | --- |
| Outcome |  | Lifestyle vs Placebo | | |
|  |  | estimate | 95% CL | p value |
| Digit Symbol Substitution Test Score | Model 1 | -0.83 | -2.2, 0.5 | 0.2 |
|  | Model 2 | -0.37 | -1.4, 0.67 | 0.5 |
|  | Model 3 | -0.37 | -1.4, 0.67 | 0.5 |
|  | Model 4 | -0.7 | -1.8, 0.41 | 0.2 |
| Spanish English Verbal Learning Test — Immediate Recall Score | Model 1 | -0.01 | -0.63, 0.61 | 0.9 |
|  | Model 2 | 0.14 | -0.35, 0.63 | 0.6 |
|  | Model 3 | 0.14 | -0.35, 0.63 | 0.6 |
|  | Model 4 | 0.13 | -0.41, 0.68 | 0.6 |
| Model 1: visit year + treatment assignment | |  |  |  |
| Model 2: Model 1+ age + sex + race + education + income category | | |  |  |
| Model 3: Model 2 + APOE Ɛ4 carrier status | |  |  |  |

**Supplemental Figure 1.** Consort diagram

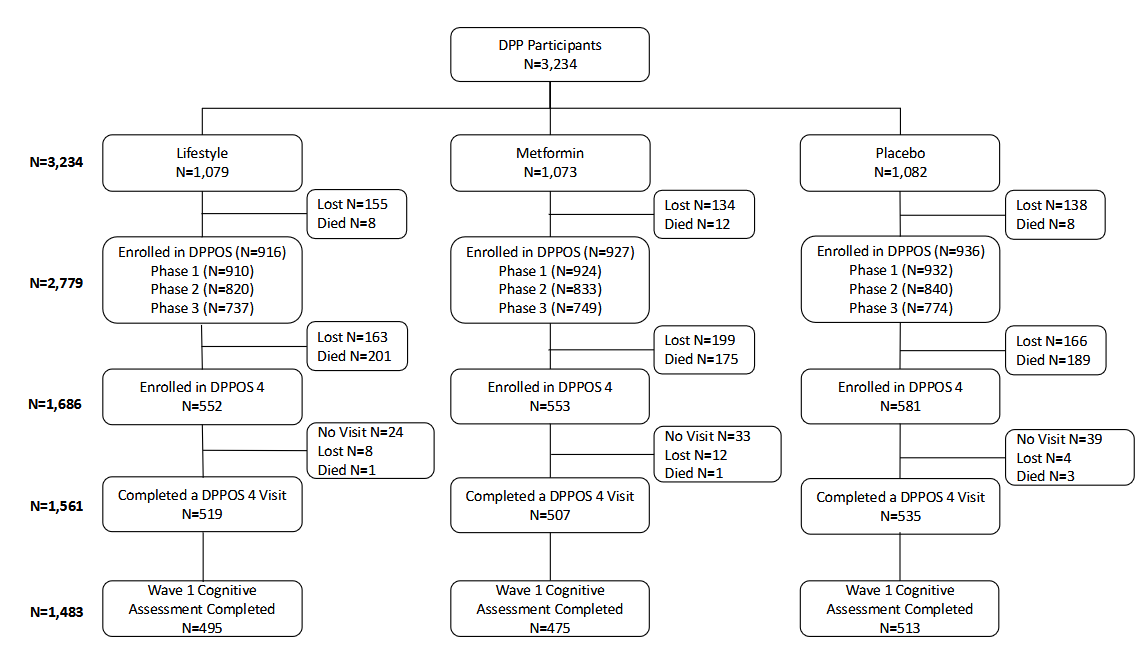

**Supplemental Figure 2.** Cumulative metformin exposure by DPP treatment assignment over the DPP/DPPOS study period, and time-specific proportions of participants using metformin (including out-of-study metformin for all three treatment groups as well as study-provided metformin in the metformin group). Study-provided metformin was masked during DPP and then provided unmasked to the original metformin group participants until 2022.

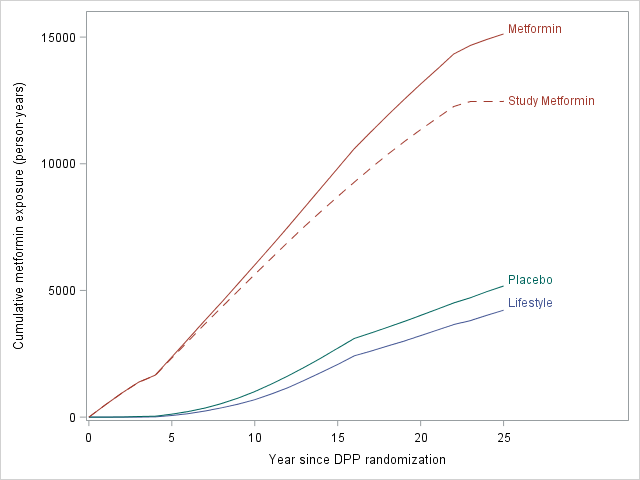

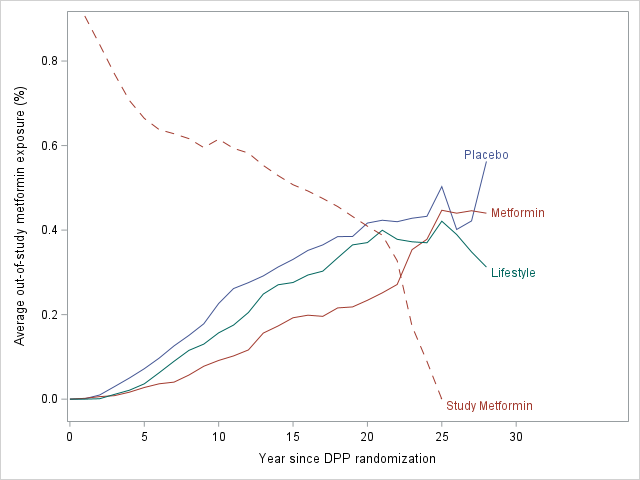
