## Supplementary material for "Randomized metformin and cognitive outcomes in the Diabetes Prevention Program Outcomes Study": DPPOS Investigators Appendix

**Pennington Biomedical Research Center**

**(Baton Rouge, LA)**

George A. Bray, MD*

Kishore M. Gadde, MD*

Owen Carmichael, PhD*

Jennifer Levatino RN, BSN**

Amber Dragg RD, LDN**

Frank Greenway, MD

Stephen Lee, BS

Donald Lewis

**University of Chicago (Chicago, IL)**

Kenneth S. Polonsky, MD*

Janet Tobian, MD, PhD*

David A. Ehrmann, MD*

Margaret J. Matulik, RN, BSN**

Karla A. Temple, PhD, RDN, LDN**

Bart Clark, MD

Kirsten Czech, MS

Catherine DeSandre, BA

Brittnie Dotson, MS

Ruthanne Hilbrich, RD

Wylie McNabb, EdD

Ann R. Semenske, MS, RD

Celeste C. Thomas, MD

Marisela Vargas, BA

**Jefferson Medical College (Philadelphia, PA)**

Jose F. Caro, MD*

Kevin Furlong, DO*

Barry J. Goldstein, MD, PhD*

Pamela G. Watson, RN, ScD*

Kellie A. Smith, RN, MSN**

Jewel Mendoza, RN, BSN**

Marsha Simmons, CCRP**

Wendi Wildman, RN**

Renee Liberoni, MPH

John Spandorfer, MD

Constance Pepe, MS, RD

Marie-France Hivert, MD

Catherine Lark

Eden Mikalic, BSN

Rudolph McClain

Genine Jensen, RN

**University of Miami (Miami, FL)**

Richard P. Donahue, PhD*

Ronald B. Goldberg, MD*

Ronald Prineas, MD, PhD*

Diana Soliman MHS, MD*

Jeanette Calles, MSEd**

Anna Giannella, RD, MS**

Patricia Rowe, MPA**

Juliet Sanguily, RN**

Hermes J. Florez, MD

Bertha Veciana

**The University of Texas Health Science Center**

**(San Antonio, TX)**

Steven M. Haffner, MD, MPH*

Helen P. Hazuda, PhD*

Maria G. Montez, RN, MSHP, CDE**

Kathy Hattaway, RD, MS

Juan Isaac, RN, BSN**

Carlos Lorenzo, MD, PhD

Arlene Martinez, RN, BSN, CDE

Monica Salazar

Tatiana Walker, RD, MS, CDE

**University of Colorado (Denver, CO)**

Dana Dabelea, MD, PhD*

Richard F. Hamman, MD, DrPH*

Lisa Testaverde, MS**

Jennifer Truong, MPH**

Thomas Nilan, BS**

Leigh Perrault, MD

Allison Shapiro, PhD, MPH

Devin Grove, BS

Lia Kopesky, BS

Maxine Kugelmas, BS

Jody Tanabe, MD

**Joslin Diabetes Center (Boston, MA)**

Edward S. Horton, MD*

Medha Munshi, MD*

Kathleen E. Lawton, RN**

Sharon D. Jackson, CCRC,MS, RD, CDE**

Catherine S. Poirier, RN, BSN**

Kati Swift, RN, BSN**

Christine Slyne, BA**

Ronald A. Arky, MD

Marybeth Bryant

Jacqueline P. Burke, BSN

Enrique Caballero, MD

Karen M. Callaphan, BA

Barbara Fargnoli, RD

Therese Franklin

Om P. Ganda, MD

Ashley Guidi, BS

Mathew Guido, BA

Alan M. Jacobsen, MD

Lyn M. Kula, RD

Margaret Kocal, RN, CDE

Lori Lambert, MS, RD, LD

Kathleen E. Lawton, RN

Sarah Ledbury, Med, RD

Maureen A. Malloy, BS

Roeland J.W. Middelbeek, MD

Maryanne Nicosia, MS, RD

Cathryn F. Oldmixon, RN

Jocelyn Pan, BS, MPH

Marizel Quitingon

Riley Rainville, BS

Stacy Rubtchinsky, BS

Ellen W. Seely, MD

Jessica Sansoucy, BS

Dana Schweizer, BSN

Donald Simonson, MD

Fannie Smith, MD

Caren G. Solomon, MD, MPH

Jeanne Spellman, RD

James Warram, MD

Bridget Carey, BS

Colin Conery, BS

**VA Puget Sound Health Care System and University of Washington (Seattle, WA)**

Steven E. Kahn, MB, ChB*

Pandora L. Wander, MD, MS*

Basma N. Fattaleh, BA **

Brenda K. Montgomery, RN, BSN, CDE**

Celeste Colegrove, BS

Wilfred Fujimoto, MD

Robert H. Knopp, MD

Edward W. Lipkin, MD

Michelle Marr, BA

Ivy Morgan-Taggart

Anne Murillo, BS

Kayla O’Neal, BS

Dace Trence, MD

Lonnese Taylor, RN, BS

April Thomas, RD, MPH, CDE

Elaine C. Tsai, MD, MPH

**University of Tennessee (Memphis, TN)**

Samuel Dagogo-Jack, MD, DSc, FRCP, FACP*

Abbas E. Kitabchi, PhD, MD, FACP*

Mary E. Murphy, RN, MS, CDE, MBA**

Laura Taylor, RN, BSN, CDE**

Jennifer Dolgoff, RN, BSN**

Ethel Faye Hampton, R.N.**

William B. Applegate, MD, MPH

Michael Bryer-Ash, MD

Debra Clark, LPN

Sandra L. Frieson, RN

Uzoma Ibebuogu, MD

Raed Imseis, MD

Helen Lambeth, RN, BSN

Lynne C. Lichtermann, RN, BSN

Hooman Oktaei, MD

Harriet Ricks

Lily M.K. Rutledge, RN, BSN

Amy R. Sherman, RD, LD

Clara M. Smith, RD, MHP, LDN

Judith E. Soberman, MD

Beverly Williams-Cleaves, MD

Avnisha Patel, MLT

Ebenezer A. Nyenwe, MD, FACP

Usman Baguda, MBBS

Samson Iwhiwhu, MBBS

**Northwestern University’s Feinberg School of Medicine (Chicago, IL)**

Boyd E. Metzger, MD*

Mark E. Molitch, MD*

Amisha Wallia, MD*

Mariana K. Johnson, MS, RN**

Christina Coventry, MS, RN**

Daphne T. Adelman, MBA, RN

Catherine Behrends

Michelle Cook, MS

Marian Fitzgibbon, PhD

Mimi M. Giles, MS, RD

Deloris Heard, MA

Cheryl K.H. Johnson, MS, RN

Diane Larsen, BS

Anne Lowe, BS

Megan Lyman, BS

David McPherson, MD

Samsam C. Penn, BA

Thomas Pitts, MD

Renee Reinhart, RN, MS

Susan Roston, RN, RD

Pamela A. Schinleber, RN, MS

Matthew O’Brien, MD

Monica Hartmuller, MS, RN

**Massachusetts General Hospital (Boston, MA)**

David M. Nathan, MD*

Camille Powe, MD*

Charles McKitrick, BSN**

Heather Turgeon, BSN**

Mary Larkin, MSN, RN, CDCES**

Lindsey Gurry, BSN, RN, CDE**

Nopporn Thangthaeng, PhD, BSN, CDCES**

Marielle Mugford, BA

Kathy Abbott

Ellen Anderson, MS, RD

Laurie Bissett, MS, RD

Kristy Bondi, BS

Enrico Cagliero, MD

Jose C. Florez, MD, PhD+

Linda Delahanty, MS, RD

Valerie Goldman, MS, RD

Elaine Grassa

Kali D’Anna

Fernelle Leandre, BS

Peter Lou, MD

Alexandra Poulos

Elyse Raymond, BS

Valerie Ripley, BS

Christine Stevens, RN

Beverly Tseng

Kathy Chu, BA

Rachel Bartholomew, BA

**University of California-San Diego (La Jolla, CA)**

Jerrold M. Olefsky, MD*

Elizabeth Barrett-Connor, MD*

Sunder Mudaliar, MD*

Maria Rosario Araneta, PhD*

Mary Lou Carrion-Petersen, RN, BSN**

Karen Vejvoda, RN, BSN, CDE, CCRC**

Sarah Bassiouni, MPH

Madeline Beltran, RN, BSN, CDE

Lauren N. Claravall, BS

Jonalle M. Dowden, BS

Steven V. Edelman, MD

Pranav Garimella, MBBS

Robert R. Henry, MD

Javiva Horne, RD

Marycie Lamkin, RN

Simona Szerdi Janesch, BA

Diana Leos

William Polonsky, PhD

Rosa Ruiz

Jean Smith, RN

Jennifer Torio-Hurley

Gabrielle Armijo, MD

Melissa Rojas-Cuevas, BS

**Columbia University (New York, NY)**

F. Xavier Pi-Sunyer, MD*

Blandine Laferrere, MD, PhD*

Jane E. Lee, MS**

Susan Hagamen, MS, RN, CDE**

Kim Kelly-Dinham**

Jose A. Luchsinger, MD, MPH

Rabiah Borhan

Julie Roman

**Indiana University (Indianapolis, IN)**

Mary de Groot, PhD*

David G. Marrero, PhD*

Kieren J. Mather, MD*

Melvin J. Prince, MD*

Susie M. Kelly, RN, CDE**

Marcia A. Jackson**

Gina McAtee**

Paula Putenney, RN**

Ronald T. Ackermann, MD

Carolyn M. Cantrell

Yolanda F. Dotson, BS

Edwin S. Fineberg, MD

Megan Fultz

John C. Guare, PhD

Angela Hadden

James M. Ignaut, MA

Marion S. Kirkman, MD

Erin O’Kelly Phillips

Kisha L Pinner

Beverly D. Porter, MSN

Paris J. Roach, MD

Nancy D. Rowland, BS, MS

Madelyn L. Wheeler, RD

Rachel Klingensmith

**Medstar Research Institute (Washington, DC)**

Vanita Aroda, MD*

Michelle Magee, MD*

Robert E. Ratner, MD*

Michelle Magee, MD*

Gretchen Youssef, RD, CDE**

Sue Shapiro, RN, BSN, CCRC**

Natalie Andon, RN

Catherine Bavido-Arrage, MS, RD, LD

Geraldine Boggs, MSN, RN

Marjorie Bronsord, MS, RD, CDE

Ernestine Brown

Holly Love Burkott, RN

Wayman W. Cheatham, MD

Susan Cola

Cindy Evans

Peggy Gibbs

Tracy Kellum, MS, RD, CDE

Lilia Leon

Milvia Lagarda

Claresa Levatan, MD

Milajurine Lindsay

Asha K. Nair, BS

Jean Park, MD

Maureen Passaro, MD

Angela Silverman

Gabriel Uwaifo, MD

Debra Wells-Thayer, NP, CDE

Renee Wiggins, RD

Raymond Turner, MD

Diana Evans

**University of Southern California/UCLA Research Center (Alhambra, CA)**

Mohammed F. Saad, MD*

Karol Watson, MD*

Anthony Sosa**

Sameh Tadros**

Preethi Srikanthan, MD

Tamara Horwich, MD

Kathy Ngo

Michelle Chan

Veronica Villarreal

**Washington University (St. Louis, MO)**

Angela L. Brown, MD*

Tamara Stich, RN, MSN, CDE**

Elizabeth Hoffmann, RN, BSN

**Johns Hopkins School of Medicine**

**(Baltimore, MD)**

Sherita Hill Golden, MD, MHS, FAHA*

Christopher D. Saudek, MD*

Vanessa Bradley, BA**

Emily Sullivan, MEd, RN**

Tracy Whittington, BS**

Caroline Abbas

Engle Abrams

Adrienne Allen

Umaima Tahir Banda, MPH

Frederick L. Brancati, MD, MHS

Sharon Cappelli

Jeanne M. Clark, MD

Jeanne B. Charleston, RN, MSN

Janice Freel

Katherine Horak, RD

Alicia Greene

Dawn Jiggetts

Deloris Johnson

Hope Joseph

Kimberly Loman

Nestoras Mathioudakis, MD, MHS

Henry Mosley

John Reusing

Richard R. Rubin, PhD

Alafia Samuels, MD

Thomas Shields

Shawne Stephens

Kerry J. Stewart, EdD

LeeLana Thomas

Evonne Utsey

Paula Williamson

**University of New Mexico (Albuquerque, NM)**

David S. Schade, MD*

Anna Korbin, BS**

Elizabeth Duran-Valdez, MS

Carolyn King, MEd

Kakin Szeto

Emma Montaño

**Albert Einstein College of Medicine (Bronx, NY)**

Jill Crandall, MD*

Sandra Aleksic, MD*

Danielle Powell, MPH**

Norica Tomuta, MD

Janet O. Brown, RN, MPH, MSN

Gilda Trandafirescu, MD

Harry Shamoon, MD

Elizabeth A. Walker PhD, RN

Judith Wylie-Rosett, EdD, RD

**University of Pittsburgh (Pittsburgh, PA)**

Trevor Orchard, MD*

Elizabeth Venditti, PhD*

Rena R. Wing, PhD*

Susan Jeffries, RN, MSN**

Gaye Koenning, MS, RD**

M. Kaye Kramer, BSN, MPH**

Marie Smith, RN, BSN**

Susan Barr, BS

Catherine Benchoff

Miriam Boraz, PhD

Lisa Clifford, BS

Rebecca Culyba, BS

Marlene Frazier

Ryan Gilligan, BS

Stephanie Guimond, BS

Susan Harrier, MLT

Louann Harris, RN

Andrea Kriska, PhD

Qurashia Manjoo, MD

Monica Mullen, MHP, RD

Alicia Noel, BS

Amy Otto, PhD

Jessica Pettigrew, CMA

Bonny Rockette-Wagner, PhD

Debra Rubinstein, MD

Linda Semler, MS, RD

Cheryl F. Smith, PhD

Valarie Weinzierl, MPH

Katherine V. Williams, MD, MPH

Tara Wilson, BA

Bonnie Gillis, MS, RD, LDN

**University of Hawaii (Honolulu, HI)**

Marjorie K. Mau, MD*

Narleen K. Baker-Ladao, BS**

John S. Melish, MD

Richard F. Arakaki, MD*

Renee W. Latimer, BSN, MPH**

Mae K. Isonaga, RD, MPH**

Ralph Beddow, MD

Nina E. Bermudez, MS

Lorna Dias, AA

Jillian Inouye, RN, PhD

Kathy Mikami, BS, RD

Pharis Mohideen, MD

Sharon K. Odom, RD, MPH

Raynette U. Perry, AA

Robin E. Yamamoto, CDE, RD

Michiko Bruno, MD

**Southwest American Indian Centers**

**(Phoenix, AZ; Shiprock, NM; Zuni, NM)**

William C. Knowler, MD, DrPH*+

Robert L. Hanson, MD, MPH*

Vallabh Raj Shah, PhD, MS, FASN*

Harelda Anderson, LMSW**

Norman Cooeyate**

Charlotte Dodge**

Mary A. Hoskin, RD, MS**

Carol A. Percy, RN, MS**

Alvera Enote**

Camille Natewa**

Kelly J. Acton, MD, MPH

Vickie L. Andre, RN, FNP

Rosalyn Barber

Miranda Smart

Sara Michaels, MD

Kevin McDermott, PharmD

Laura Urbanski, MD

Shandiin Begay, MPH

Peter H. Bennett, MB, FRCP

Mary Beth Benson, RN, BSN

Evelyn C. Bird, RD, MPH

Brenda A. Broussard, RD, MPH, MBA, CDE

Brian C. Bucca, OD, FAAO

Marcella Chavez, RN, AS

Sherron Cook

Jeff Curtis, MD

Tara Dacawyma

Matthew S. Doughty, MD

Roberta Duncan, RD

Cyndy Edgerton, RD

Jacqueline M. Ghahate

Justin Glass, MD

Martia Glass, MD

Dorothy Gohdes, MD

Wendy Grant, MD

Robert L. Hanson, MD, MPH

Ellie Horse

Louise E. Ingraham, MS, RD, LN

Merry Jackson

Priscilla Jay

Roylen S. Kaskalla

Karen Kavena, ANP

David Kessler, MD

Kathleen M. Kobus, RNC-ANP

Jonathan Krakoff, MD

Jason Kurland, MD

Catherine Manus, LPN

Cherie McCabe

Sara Michaels, MD

Tina Morgan

Yolanda Nashboo

Julie A. Nelson, RD

Steven Poirier, MD

Evette Polczynski, MD

Christopher Piromalli, DO

Mike Reidy, MD

Jeanine Roumain, MD, MPH

Debra Rowse, MD

Robert J. Roy

Sandra Sangster, RD

Janet Sewenemewa

Miranda Smart

Chelsea Spencer

Darryl Tonemah, PhD

Rachel Williams, FNP

Charlton Wilson, MD

Michelle Yazzie

Alphonse Alapat RN, MSN, ACNP-AG, CNN

**George Washington University Biostatistics Center (DPP Coordinating Center Rockville, MD)**

Raymond Bain, PhD*

Sarah Fowler, PhD*

Marinella Temprosa, PhD*

Michael D. Larsen, PhD*

Kathleen Jablonski, PhD*

Tina Brenneman**

Sharon L. Edelstein, ScM**

Solome Abebe, MS

Julie Bamdad, MS

Melanie Barkalow

Joel Bethepu

Tsedenia Bezabeh

Anna Bowers

Nicole Butler

Jackie Callaghan

Caitlin E. Carter

Costas Christophi, PhD

Gregory M. Dwyer, MPH

Mary Foulkes, PhD

Yuping Gao

Robert Gooding

Adrienne Gottlieb

Kristina L. Grimes

Nisha Grover-Fairchild, MPH

Lori Haffner, MS

Heather Hoffman, PhD

Steve Jones

Tara L. Jones

Richard Katz, MD

Preethy Kolinjivadi, MS

John M. Lachin, ScD

Yong Ma, PhD

Pamela Mucik

Robert Orlosky

Qing Pan, PhD

Susan Reamer

James Rochon, PhD

Alla Sapozhnikova

Hanna Sherif, MS

Charlotte Stimpson

Ashley Hogan Tjaden, MPH

Fredricka Walker-Murray

Audrey McMaster

Rhea Mundra

Hannah Rapoport

Nolan Kuenster

**Lifestyle Resource Core**

Elizabeth M. Venditti, PhD*

Andrea M. Kriska, PhD

Linda Semler, MS, RD, LDN

Valerie Weinzierl, MPH

**Central Biochemistry Laboratory (Seattle, WA)**

Santica Marcovina, PhD, ScD*

F. Alan Aldrich**

Jessica Harting**

John Albers, PhD

Greg Strylewicz, PhD

**Central Biochemistry Laboratory (Minneapolis, MN)**

Robert Janicek, MT, CLS*

Anthony Killeen, MD, PhD

Deanna Gabrielson, MLS (ASCP)^CM^, PMP

**NIH/NIDDK (Bethesda, MD)**

R. Eastman, MD

Judith Fradkin, MD

Sanford Garfield, PhD

Christine Lee, MD, MS

**Centers for Disease Control & Prevention**

**(Atlanta, GA)**

Edward Gregg, PhD

Ping Zhang, PhD

**Carotid Ultrasound**

Dan O’Leary, MD*

Gregory Evans

**Coronary Artery Calcification Reading Center**

Matthew Budoff, MD

Chris Dailing

**CT Scan Reading Center**

Elizabeth Stamm, MD*

**Dual Energy X-ray Absorptiometry Reading Center (San Francisco, CA)**

Ann Schwartz, PhD

Caroline Navy

Lisa Palermo, MS

**Epidemiological Cardiology Research Center- Epicare (Winston-Salem, NC)**

Pentti Rautaharju, MD, PhD*

Ronald J. Prineas, MD, PhD**

Teresa Alexander

Charles Campbell, MS

Sharon Hall

Yabing Li, MD

Margaret Mills

Nancy Pemberton, MS

Farida Rautaharju, PhD

Zhuming Zhang, MD

Elsayed Z. Soliman, MD*

Julie Hu, MSc

Susan Hensley, BS

Lisa Keasler

Tonya Taylor

**Fundus Photo Reading Center (Madison, WI)**

Barbara Blodi, MD*

Ronald Danis, MD*

Matthew Davis, MD*

Larry Hubbard*

Ryan Endres**

Deborah Elsas**

Samantha Johnson**

Dawn Myers**

Nancy Barrett

Heather Baumhauer

Wendy Benz

Holly Cohn

Ellie Corkery

Kristi Dohm

Amitha Domalpally, MD, PhD

Vonnie Gama

Anne Goulding

Andy Ewen

Cynthia Hurtenbach

Daniel Lawrence

Kyle McDaniel

Jeong Pak

James Reimers

Ruth Shaw

Maria Swift

Pamela Vargo, CRA

Sheila Watson

**Neurocognitive Assessment Group**

Jose A. Luchsinger, MD, MPH*

Danurys Sanchez, MS

James M. Noble, MD, MS

Terry Goldberg, PhD

Neelesh Nadkarni, MD PhD

Dianilka Martinez, MPH

**Nutrition Coding Center (Columbia, SC)**

Elizabeth Mayer-Davis, PhD*

Robert R. Moran, PhD**

**Quality of Well-Being Center (La Jolla, CA)**

Ted Ganiats, MD*

Kristin David, MHP*

Andrew J. Sarkin, PhD*

Erik Groessl, PhD

Naomi Katzir

Helen Chong, MA

**University of Michigan (Ann Arbor, MI)**

William H. Herman, MD, MPH

Michael Brändle, MD, MS

Morton B. Brown, PhD

Stanley Kuo, PhD

**Neuroimaging Reading Center (Philadelphia, PA)**

Ilya Nasrallah, MD, PhD

Lisa Desiderio, RT (R) (MR) CCRC

Leeanne Lezotte

Kristina Vineis

**+Genetics Working Group**

Jose C. Florez, MD, PhD^1, 2^

David Altshuler, MD, PhD^1, 2^

Liana K. Billings, MD^1^

Ling Chen, MS^1^

Maegan Harden, BS^2^

Robert L. Hanson, MD, MPH^3^

William C. Knowler, MD, DrPH^3^

Toni I. Pollin, PhD^4^

Alan R. Shuldiner, MD^4^

Kathleen Jablonski, PhD^5^

Paul W. Franks, PhD, MPhil, MS^6, 7, 8^

Marie-France Hivert, MD^8^

1=Massachusetts General Hospital

2=Broad Institute
